# Wastewater Monitoring of SARS-CoV-2 from Acute Care Hospitals Identifies Nosocomial Transmission and Outbreaks

**DOI:** 10.1101/2021.02.20.21251520

**Authors:** Nicole Acosta, María A. Bautista, Jordan Hollman, Janine McCalder, Alexander Buchner Beaudet, Lawrence Man, Barbara J. Waddell, Jianwei Chen, Carmen Li, Darina Kuzma, Srijak Bhatnagar, Jenine Leal, Jon Meddings, Jia Hu, Jason L. Cabaj, Norma J. Ruecker, Christopher Naugler, Dylan R. Pillai, Gopal Achari, M. Cathryn Ryan, John M. Conly, Kevin Frankowski, Casey RJ Hubert, Michael D. Parkins

## Abstract

**Background:** SARS-CoV-2 has been detected in wastewater and its abundance correlated with community COVID-19 cases, hospitalizations and deaths. We sought to use wastewater-based detection of SARS-CoV-2 to assess the epidemiology of SARS-CoV-2 in hospitals.

**Methods:** Between August and December 2020, twice-weekly wastewater samples from three tertiary-care hospitals (totaling >2100 dedicated inpatient beds) were collected. Wastewater samples were concentrated and cleaned using the 4S-silica column method and assessed for SARS-CoV-2 gene-targets (N1, N2 and E) and controls using RT-qPCR. Wastewater SARS-CoV-2 as measured by quantification cycle (Cq), genome copies and genomes normalized to the fecal biomarker PMMoV were compared to the total daily number of patients hospitalized with active COVID-19, confirmed cases of hospital-acquired infection, and the occurrence of unit-specific outbreaks.

**Results:** Of 165 wastewater samples collected, 159 (96%) were assayable. The N1-gene from SARS-CoV-2 was detected in 64.1% of samples, N2 in 49.7% and E in 10%. N1 and N2 in wastewater increased over time both in terms of amount of detectable virus and the proportion of samples that were positive, consistent with increasing hospitalizations (Pearson’s r=0.679, P<0.0001, Pearson’s r=0.728, P<0.0001, respectively). Despite increasing hospitalizations through the study period, wastewater analysis was able to identify incident nosocomial-acquired cases of COVID-19 (Pearson’s r =0.389, P<0.001) and unit-specific outbreaks by increases in detectable SARS-CoV-2 N1-RNA (median 112 copies/ml) versus outbreak-free periods (0 copies/ml; P<0.0001).

**Conclusions:** Wastewater-based monitoring of SARS-CoV-2 represents a promising tool for SARS-CoV-2 passive surveillance and case identification, containment, and mitigation in acute-care medical facilities.

**Supplemental Material included:** *Key-points summary:* SAS-CoV-2 RNA is detectable in hospital wastewater. Wastewater SARS-CoV-2 RNA increases in conjunction with COVID-19-related hospitalizations. Spikes in SARS-CoV-2 wastewater signal correspond to incident hospital-acquired cases and outbreaks, suggesting passive surveillance via wastewater has great promise for COVID-19 monitoring.

## INTRODUCTION

SARS-CoV-2 RNA is present in the feces of infected individuals – appearing just prior or concomitant with symptoms[1]. Accordingly, leaders in the field of wastewater-based epidemiology leveraged their expertise to study this emerging infectious disease[2, 3]. Medema *et al* first reported SARS-CoV-2-RNA in Dutch wastewater-treatment plants (WW-TP)[4]. Several groups have since adapted this technology to understand community disease-burden[5–8]. Recent studies suggest that SARS-CoV-2-RNA increases in WW-TP precede clinically diagnosed cases by 0-2 days and associated hospitalizations by 1-4 days[6].

Between 4-8% of individuals with COVID-19 will be hospitalized, with age and co-morbidities being key risk factors[9–11]. Nosocomial-transmission and outbreaks affecting patients and health care workers (HCW) are not uncommon, although drivers remain to be fully understood[12, 13]. While hospital-acquisition is rare (0.8-5 cases/10,000 patient-days in communities with high disease burden), public fear of acquiring COVID-19 from hospitals has resulted in reduced health-resource utilization and hospital avoidance, often to the detriment of patients[14, 15]. Accordingly, hospital-based detection tools are needed to understand the epidemiology of COVID-19 and potentially mitigate spread.

Hospitals hold great promise in understanding SARS-CoV-2 wastewater-generated data. Owing to their proximity to affected individuals in the municipal sewershed relative to WW-TP (i.e., shorter transit time for signal degradation[1]), hospitals may aid in understanding SARS-CoV-2 wastewater dynamics. Compared to the general community, hospitals are much more likely to comprehensively monitor and identify all cases within their populations. Furthermore, outbreaks in hospitals are rapidly and comprehensively investigated. For these reasons we embarked on this study to determine relationships between hospital SARS-CoV-2 wastewater dynamics and COVID-19 hospitalizations, nosocomial-transmissions and outbreaks.

## METHODS

### Acute-care hospitals and hospital information systems

We monitored SARS-CoV-2-RNA in the wastewater from three of Calgary’s four adult tertiary-care hospitals, accounting for 89% of staffed-inpatient beds (Supplementary Material). Daily prevalent-hospitalized cases were defined as those with laboratory-confirmed COVID-19 within 14 days, remaining on contact/droplet precautions. Hospital-acquired cases were defined as patients who were admitted to hospital ≥7 days before COVID-19 symptom onset that were then confirmed by a positive RT-qPCR SARS-CoV-2 test; or a patient admitted to hospital for ≤7 days confirmed to have hospital-acquired COVID-19 infection based on an epidemiological link. Hospital-acquired cases were separately recorded including the unit where they were acquired and are reported as hospital-wide signals for Hospital-1 and 2. Data for Hospital-3 are presented as 3A, 3B, 3C based on wastewater drainage outflows of different units. COVID-19 outbreaks were defined as any unit with ≥1 confirmed hospital-acquired case(s) and/or ≥2 confirmed COVID-19 cases in HCWs linked to a unit. This research was approved by the University of Calgary’s Conjoint Health Regional Ethics Board (REB-20-1252).

### Wastewater sampling

Wastewater samples were collected from August 5^th^ to December 17^th^, 2020 at three hospitals. Hospital-wide access through a single sampling point was not possible at Hospital-3 where an initial sampling point (Hospital-3A) captured the units predicted to be most relevant for COVID-19 (including ICUs and dedicated COVID-19 care-units). Beginning October 1^st^ two additional sites were added, Hospital-3B and Hospital-3C, expanding coverage to all inpatient-care buildings. Composite 24-hour samples of wastewater were collected using ISCO-GLS autosamplers (Lincoln, Nebraska) placed inside sewer access points outside of hospitals. Samples were taken from each location twice-weekly.

### Sample Processing and RNA Purification

Sample preparation and molecular analysis were performed at separate sites to prevent cross contamination. At ACWA, wastewater samples were processed with a 40 ml aliquot taken to comprise the sample. Each sample was spiked with a positive control - Bovilis® attenuated- Coronavirus Vaccine (BCoV; Merck, #151921). Sample processing and RNA purification was conducted using the 4S-silica column method (i.e., Sewage, Salt, Silica and SARS-CoV-2) with modifications[16]. Purified nucleic acids were transported on dry ice to the Health Sciences Centre for molecular analysis. An extraction blank control was included for every processed sample batch to ensure no contamination occurred.

### RT-qPCR analysis

We used RT-qPCR to quantify SARS-CoV-2-RNA and controls in wastewater. Specific primers and probes (Supplementary Table-S1) were used to amplify two regions of the nucleocapsid gene (i.e., N1 and N2) and a region of the envelope gene (i.e., E). Samples were considered positive for the presence of SARS-CoV-2 RNA-target if amplification passed a detection cycle threshold in <40 cycles for at least one of N1, N2 and/or E [4, 17, 18]. The quantification cycle (Cq) value was used for calculations when this threshold was <40 cycles or reached between 40 and 45 cycles for one target as long as another target reached the threshold in <40 cycles. Amplification of the Pepper mild mottle virus (PMMoV) was employed to incorporate a human fecal biomarker control in order to normalize SARS-CoV-2 for the relative bioburden in samples[5]. RT-qPCR was performed using a QuantStudio-5 Real-Time PCR System (Applied Biosystems) with each run including no-template controls (NTCs) in triplicate. For N1, N2 and E-assays three positive controls were included in each run. Samples where minimal signal for BCoV or PMMoV controls were recovered were excluded from the analysis to mitigate false-negative results. Amplified products from wastewater were also Sanger sequenced to confirm amplicons were derived from SARS-CoV-2.

### Statistical analysis

To compare the surrogate BCoV and PMMoV signals between hospitals, pairwise Mann-Whitney tests were performed. Non-parametric Kruskal-Wallis tests were performed for multiple comparisons of assay sensitivity and surrogate organism signals between sampling locations. Pearson’s correlation analyses were performed to determine the correlation of wastewater RNA-signal measured as *i)*. Cq, or *ii)*. genome copies/ml of wastewater or *iii)*. genome copies/genome of PMMoV, vs daily-hospitalized cases. To assess for correlation of wastewater RNA-signal with incident hospital-acquired cases, and to compensate for gaps owing to the twice-weekly sampling, incident cases occurring +/-3 days were compared to wastewater signals. To compare the SARS-CoV-2 wastewater N1-signal observed during and between unit-outbreaks, each sample was dichotomized as being collected within 3-days of a declared outbreak or not. Samples after a declared outbreak were excluded until 3-days after the last in-hospital linked case was identified. Statistical tests analyzed Hospital-1 and Hospital-2 together (given their capture of the entire hospital-facility) and separately. Statistical analyses were conducted with GraphPad’s Prism-8 software (La Jolla, CA).

## RESULTS

### Hospital SARS-CoV-2 Wastewater-RNA Kinetics

In total, 165 hospital wastewater samples were collected and 159 assessed through 135-days of bi-weekly observation (40 Hospital-1; 39 Hospital-2; 34 Hospital-3A; 23 Hospital-3B; 23 Hospital-3C). Six samples were excluded (Supplementary Figures S1-S2) SARS-CoV-2-RNA in wastewater increased over time in both the amount detectable and the proportion of samples that were positive, consistent with increasing cases and hospitalizations (Table 1, Supplementary Figure S3), coinciding with Calgary’s COVID-19 ‘second wave’. We observed that the N1-assay had the best sensitivity for detecting SARS-CoV-2. Of the 96 samples tested using N1, N2 and E-assays (i.e., those received between August 1^st^ and October 29^th^), 9 samples were positive for all three targets, 28 were positive only for N1 and N2, and 39 were positive only for N1. After October 29^th^ the E-assay was dropped, and 69 samples were analyzed, 51 were positive for both N1 and N2 and 63 were positive just with N1. N1 and N2-signals measured as Cq were positively correlated (Pearson’s r=0.710 Hospital-1, 0.762 Hospital-2, 0.792 Hospital-3A, 0.417 Hospital-3B and 0.491 Hospital-3C) across all sites (Table 2). No-template and blank controls for sample processing and RNA purification were negative for all assays (i.e., N1, N2, E, BCoV and PMMoV). Standard curves for all RT-qPCR assays were within an acceptable range for efficiencies (75 to 162%) and R^2^ (0.7 to 1.0) (Supplementary Table-S2).

**Table 1.**
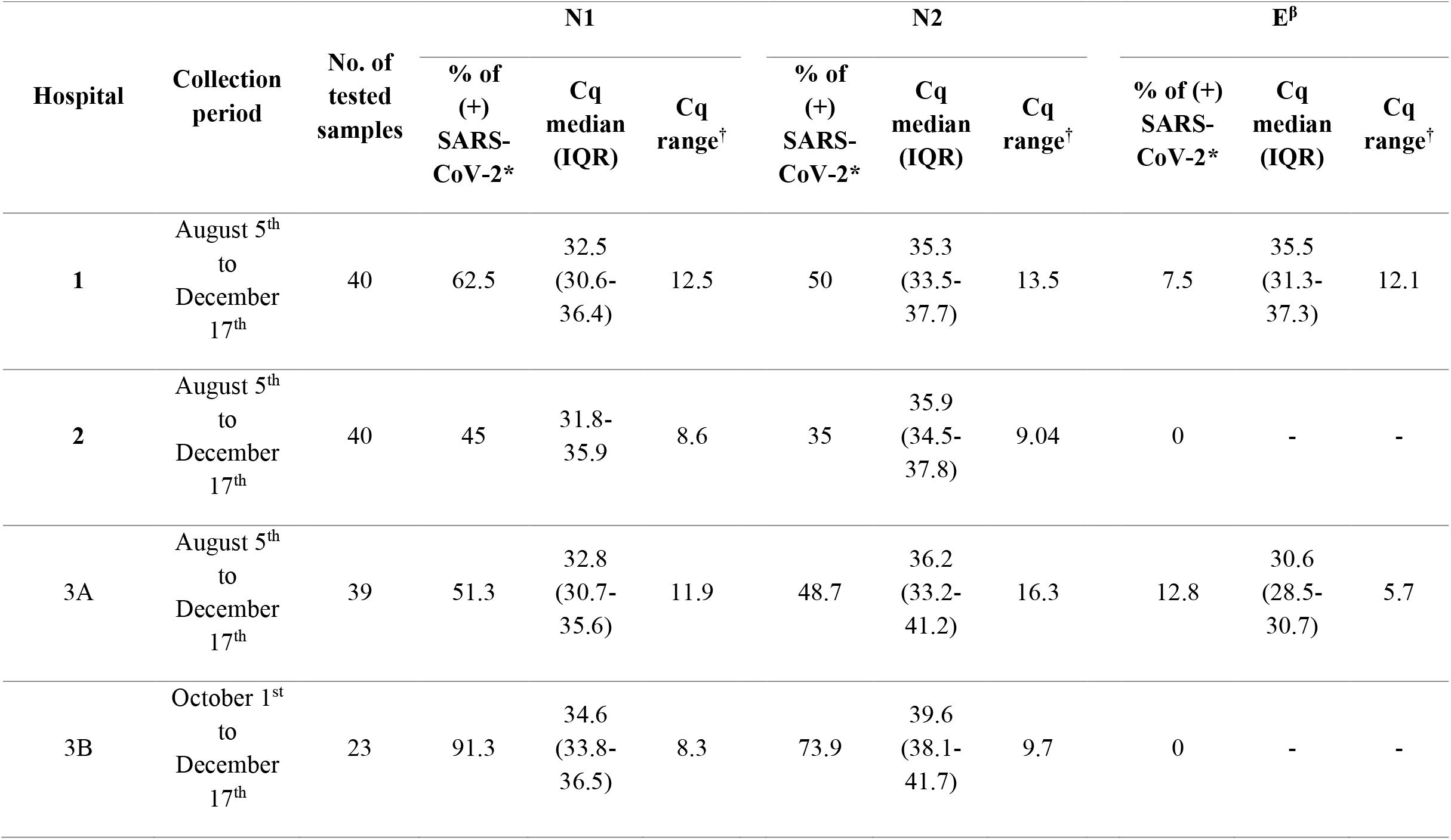
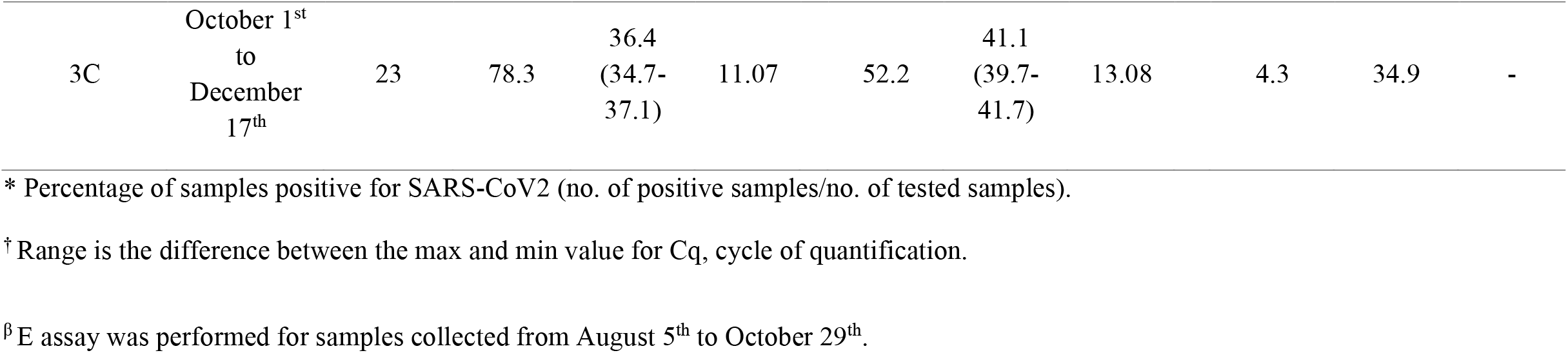
Descriptive statistics of SARS-CoV-2 RNA monitoring among wastewater hospital samples.

**Table 2.**
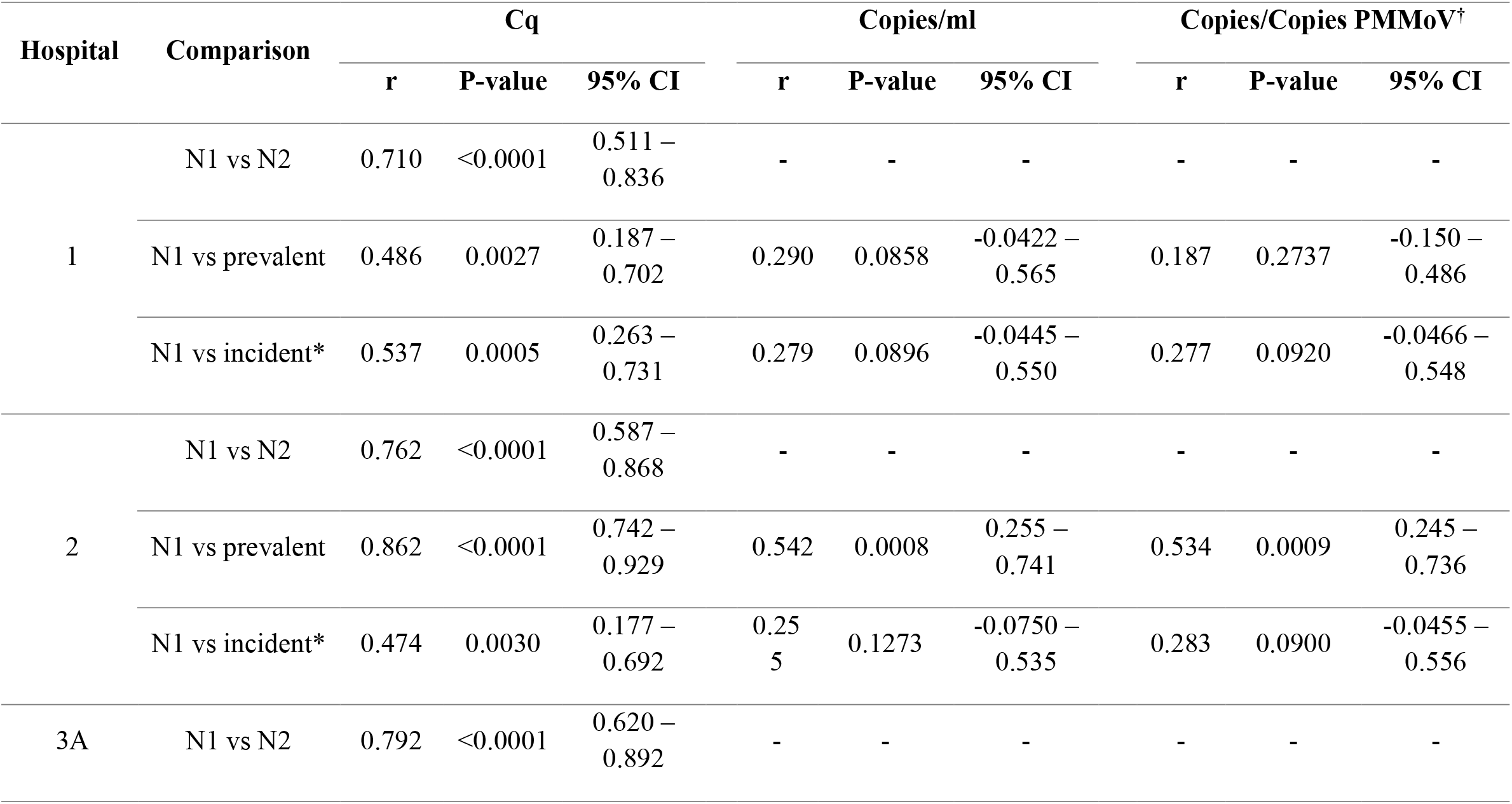
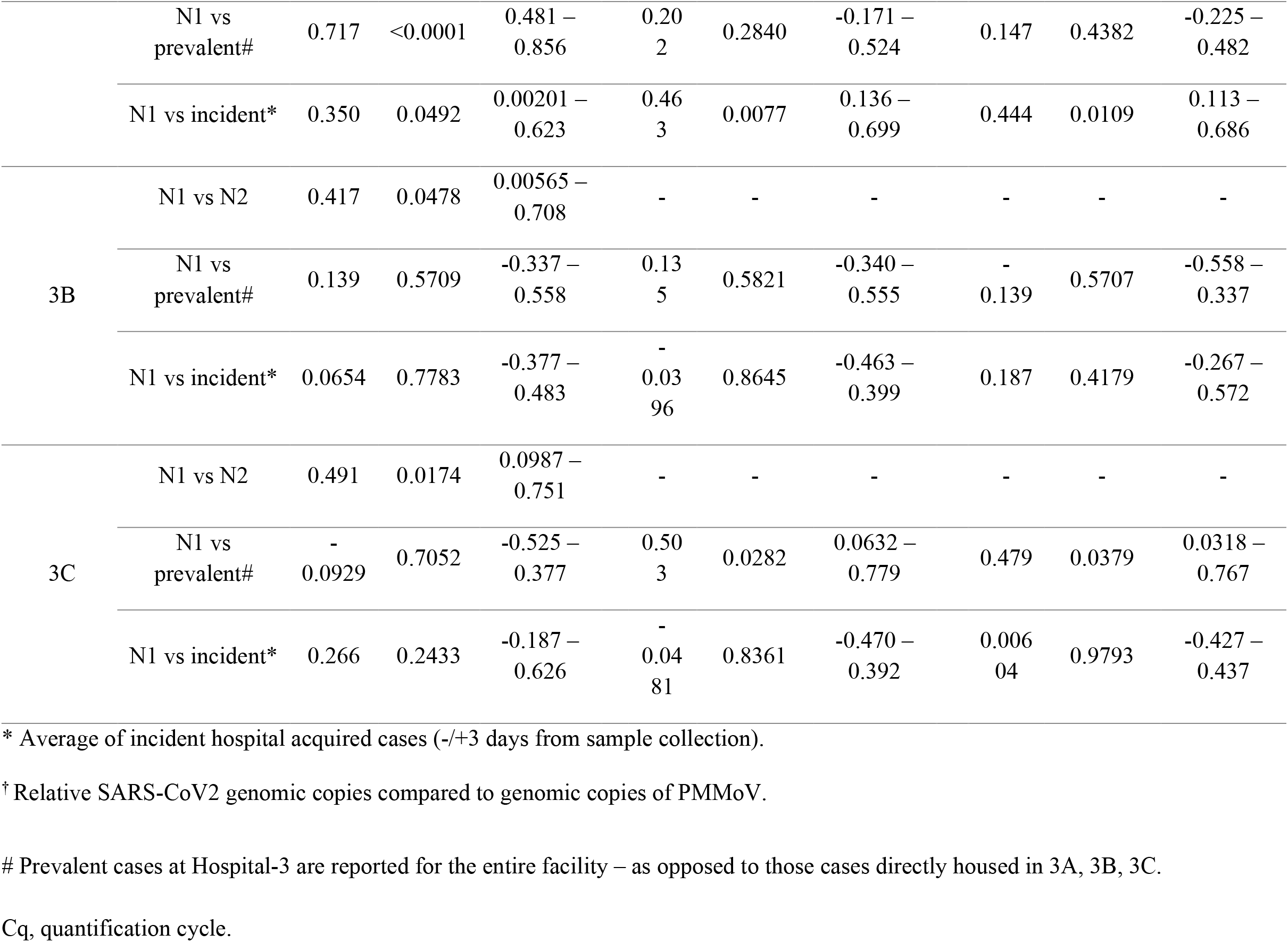
Correlation analyses between SARS-CoV-2 RNA signal in wastewater from Calgary Hospitals with daily-prevalent cases of patients hospitalized with COVID-19 and those incident hospital-acquired cases.

Hospital-1 had a higher proportion of SARS-COV-2-positive wastewater compared to Hospital-2 and Hospital-3, consistent with the higher burden of disease in NE Calgary (Table 1 and Figure 1). Following a large outbreak in Hospital-3 involving 45 patients, 43 HCW and 5 visitors (beginning in a ward not monitored via Hospital-3A site and compounded by affected patients being transferred into different units through the hospital) that was declared on September 17^th^, wastewater sampling was expanded to include additional sites; Hospital-3B and Hospital-3C (Figure 2) to enable complete capture of Hospital-3.

**Figure 1.**
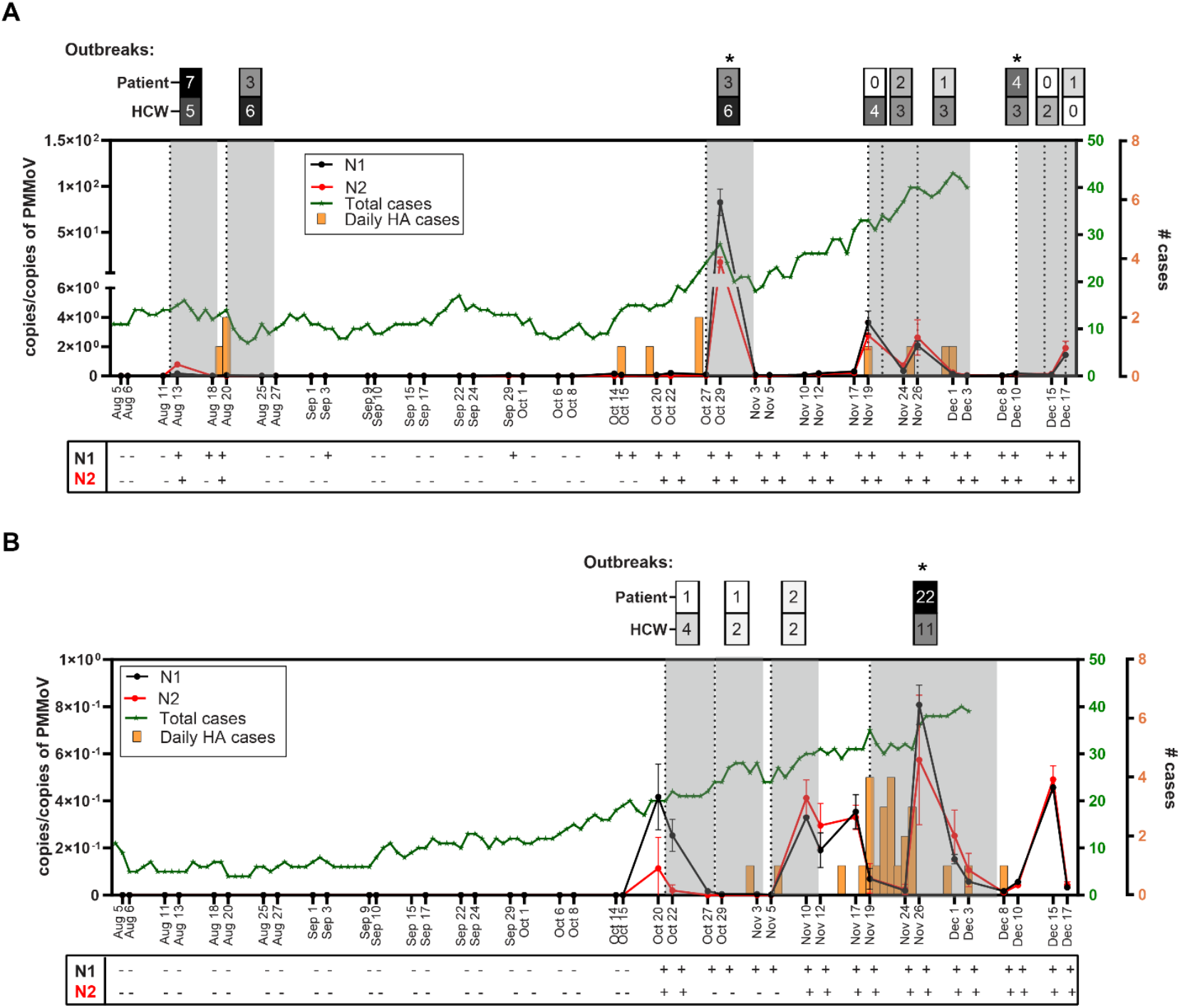
Determination of SARS-CoV-2 RNA in wastewater samples from Hospital-1 and Hospital-2. Relative SARS-CoV-2 genomic copies compared to genomic copies of PMMoV from A) Hospital 1 (August 5^th^ to December 17^th^) and B) Hospital 2 (August 5^th^ to December 17^th^). Quantification of SARS-CoV-2 RNA in samples was determined by the N1 (black) and N2 (red) assays. Green line denotes the total daily number of active prevalent cases in the hospital. Orange bars denotes the number of daily hospital-acquired cases. Plots show the average of three technical replicates and error bars represent the standard deviation. Vertical dash lines correspond to days where outbreaks were declared (Table S3), where the number of patients and health care workers involved are indicated at the top each dotted dash line. Asterisk denotes that for a specific outbreak more than one unit was involved. Gray zones denote duration of the outbreak. HA: hospital acquired. Bottom individual boxed areas represent individual samples as Positives (+) = samples where SARS-CoV-2 signal identified with a Cq<40, and negatives (-) had values ≥40. Please note that the scale is different in Figure A and B.

**Figure 2.**
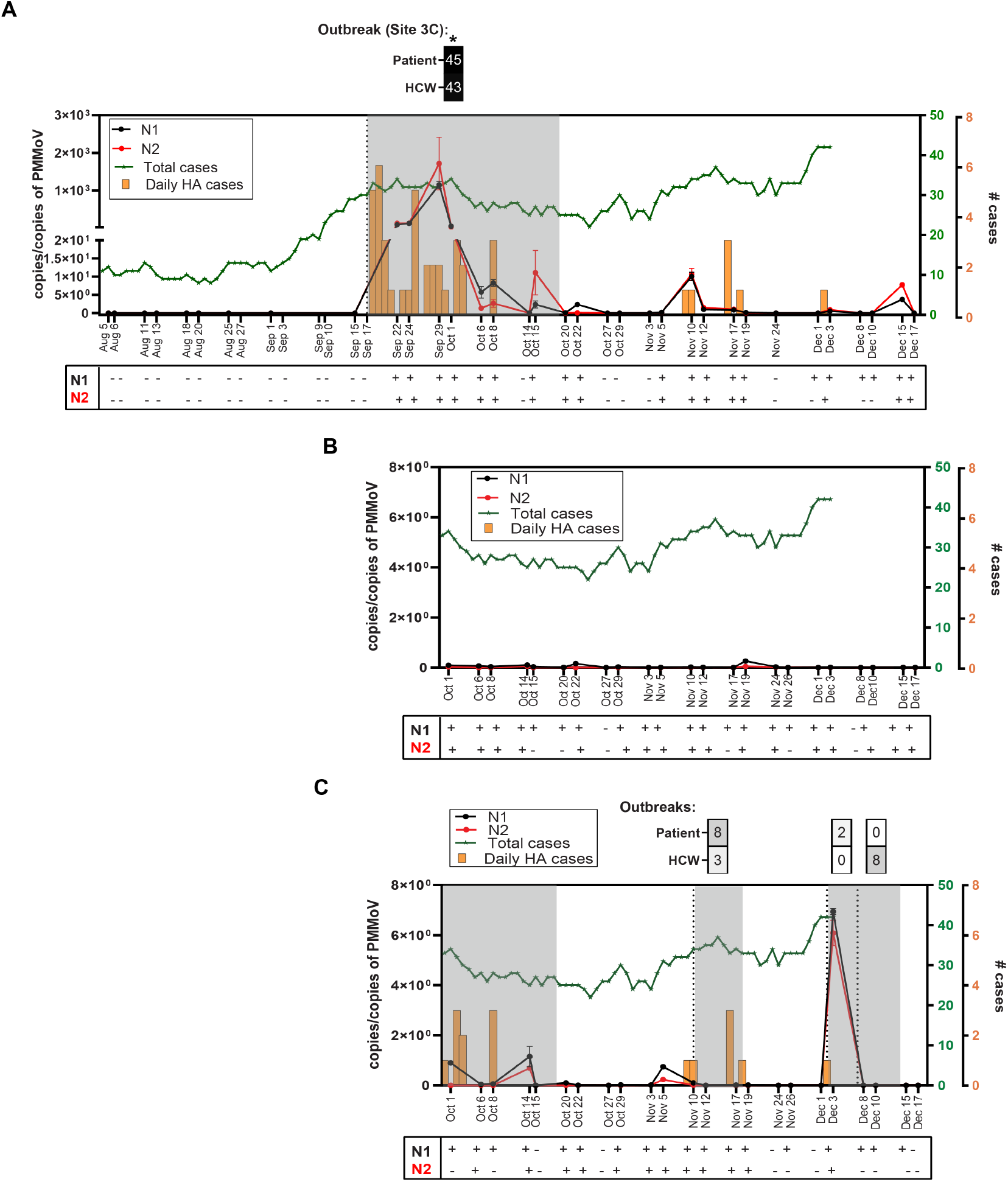
Determination of SARS-CoV-2 RNA in wastewater samples from Hospital-3. Relative SARS-CoV-2 genomic copies compared to genomic copies of PMMoV from A) Hospital 3A (Trauma, Medical & Surgical ICUs, orthopedics surgery and designated COVID-care units) August 5^th^ to December 17^th^), B) Hospital 3B (i.e., Main Building, North wing, October 1^st^ to December 17^th^) and C) Hospital3C (i.e., Main Building South Wing, cancer care building, complex medical care building and hostel/administration building), October 1^st^ to December 17^th^). Quantification of SARS-CoV-2 RNA in samples was determined by the N1 (black) and N2 (red) assays. Green line denotes the number of prevalent cases in the hospital. Orange bars denotes the number of daily hospital-acquired cases. Plots show the average of three technical replicates and error bars represent the standard deviation. Vertical dash lines correspond to days where outbreaks were declared (Table S3), where the number of patients and health care workers involved are indicated at the top each dotted dash line. Asterisk (*) denotes the largest outbreak which occurred initially at Hospital_3C prior to instituted monitoring at that site – and reflects the SARS-CoV-2 infected patients who were relocated to the designated COVID-19 wards inHospital_3A where it was detected by wastewater monitoring. The last case associated with the large outbreak was identified October 19^th^. Gray zones denote duration of the outbreak. HA: hospital acquired. Bottom individual boxed areas represent individual samples as Positives (+) = samples where SARS-CoV-2 signal identified with a Cq<40, and negatives (-) had values ≥40. Please note that the scale is different from A, B and C figures.

### Wastewater SARS-CoV-2 signal correlates with total hospitalized COVID-19 cases

We assessed the correlation between the SARS-CoV-2 wastewater-N1 with active-COVID-19 patients on contact/droplet isolation at each hospital. When assessed together, Hospital-1 and Hospital-2, we observed that as prevalent cases increased, the wastewater-signal measured as N1-Cq also increased (Pearson’s r=0.679, CI: 0.529-0.787, P<0.0001). This was also true when Hospital-1 and Hospital-2 were assessed separately (Table 2). The same was observed when SARS-CoV-2-N1 wastewater was normalized against copies of the PMMoV at Hospital-2, but only trended towards significance at Hospital-1 (Table 2). These same correlations are not as reliable at the Hospital-3 as we did not have access to prevalent cases as a function of sampling site. However, we continued to observe a positive correlation (Table 2) between prevalent cases vs N1-wastewater signal was measured as Cq at Hospital-3A (including dedicated COVID-19 care units and ICUs) (Pearson’s r=0.717) or measured as copies/ml and copies normalized to PMMoV at Hospital-3C (Pearson’s r=0.503 and 0.479, respectively).

### Wastewater SARS-COV2 signal correlates with hospital-acquired infections and outbreaks

We observed a positive correlation between wastewater N1-signal and hospital-acquired cases at Hospital-1 and Hospital-2 when analyzed together (Pearson’s r=0.389, CI: 0.177–0.566, P<0.001) and individually (Table 2). Hospital-3 data could not be fully analyzed as we did not have complete access to patient/HCW movements. Total SARS-CoV-2 as measured by Cq correlated with incident hospital-acquired cases at Hospital-3A when normalized relative to PMMoV. With respect to whether peaks in SARS-CoV-2 in wastewater associated with outbreaks, we compared SARS-CoV-2 signal from wastewater samples collected within 3 days of an outbreak being declared with samples collected during outbreak-free periods. When Hospital-1 and Hospital-2 were analysed together, we observed significant differences in median SARS-CoV-2 N-1 between outbreak-free periods vs outbreak periods when measured as copies/ml (0 [IQR: 0-57] vs 112 [IQR: 12-1726], P<0.0001) and normalized for PMMoV (0 [IQR: 0-0.06] vs 0.17 [IQR: 0.06-0.89], P=0.0003). We observed that at each of Hospital-1, Hospital-2 and Hospital-3A there were significant differences in median SARS-CoV-2-N1 measured using copies/ml between outbreak and outbreak-free periods (Table 3). Similarly, the same trend was observed at Hospital-1 and Hospital-3A when wastewater SARS-CoV-2-N1 was normalized for PMMoV (Table 3).

**Table 3.** SARS-CoV-2 RNA detection in hospital-wastewater samples as a function of proximity to a declared outbreak.

| Hospital | Measurement | Outbreak-free periods vs<br>outbreaks | P-value |
| --- | --- | --- | --- |
|  |  | (Median (IQR)) |  |
| 1 | Copies/ml | 0 (0-7.9) vs 280 (35-1768) | <0.0001 |
|  | Copies/Copies PMMoV | 0 (0-0.07) vs 0.18 (0.09-1.8) | 0.0001 |
| 2 | Copies/ml | 0 (0-0) vs 8 (1.1-17) | 0.022 |
|  | Copies/Copies PMMoV | 0 (0-0) vs 0.035 (0-0.21) | 0.453 |
| 3A | Copies/ml | 0 (0-12) vs 122 (5.9-408) | 0.031 |
|  | Copies/Copies PMMoV | 0 (0-0.11) vs 0.5 (0.01-10) | 0.026 |
| 3C | Copies/ml | 2.4 (0-8.8) vs 26 (1.1-3179) | 0.212 |
|  | Copies/Copies PMMoV | 0.013 (0-0.08) vs 0.1 (0.0003-6.9) | 0.253 |

## DISCUSSION

Hospital-associated outbreaks of COVID-19 are increasingly being reported. Early data suggests that patients with nosocomial COVID-19 may fare worse than those with community-acquired disease, experiencing longer hospital stays but not increased mortality[13]. This observation balances the opposing impacts of increased co-morbidities and medical acuity in hospitalized-individuals on one-hand, with the potential for earlier detection and more rapid supportive care/treatment on the other.

Preventing nosocomial transmission of COVID-19 is challenging for a myriad of reasons[19]. In addition to its highly infectious nature, accurate identification, triage, and effective isolation of cases is exceedingly difficult. While COVID-19 has a typical incubation period of 5-7 days, it can take as long as 14 days to manifest such that identifying evolving symptoms in previously admitted patients is challenging[20, 21]. Furthermore, up to 40% of individuals (including patients and HCW) may be asymptomatic, pauci-symptomatic, or pre-symptomatic– each just as likely to transmit infection as symptomatic individuals[22–24]. Despite rigorous infection control protocols, nosocomial infections continue to occur. Novel strategies to understand the epidemiology of SARS-CoV-2 in hospitals are therefore urgently required. One such strategy may be the monitoring of hospital wastewater[25].

To date, most wastewater-based SARS-CoV-2 RNA surveillance has focused on monitoring community burden of disease by sampling WW-TP[5–8]. More recently, moving sampling ‘upstream’ in the wastewater-network, closer to patients, is actively being explored. The most granular data comes from single-facility assessments. Passive wastewater surveillance could hold promise as an early warning strategy, adaptable to both low- and high-risk facilities. Importantly, if an incipient signal is detected in facility-wide wastewater samples, in-building plumbing systems can be strategically sampled in a nested manner in order to confirm an outbreak location.

Here we demonstrate that both the frequency of positive samples and the abundance of SARS-CoV-2 RNA in hospital wastewater systems correlated with increasing hospitalised cases – analogous to WW-TP levels correlating with the COVID-19 community-diagnosed cases[5–8]. This was most evident using raw SARS-CoV-2 Cq values but was also evident when normalized against PMMoV levels. We observed the N1-region of the nucleocapsid gene to be more sensitive than N-2, and E so low as to be dropped from our protocol. Other groups have reported similar trends in that the N1-target is the most sensitive marker in WW-TP studies[4] and cruise ships[26].

Despite nosocomial cases and outbreaks representing a small fraction of the overall population of patients hospitalized with COVID-19, these events were discernable by wastewater testing. The natural history of SARS-CoV-2-RNA presence in the gastrointestinal tract remains incompletely understood[2, 3]. Changes in fecal viral loads over the course of the disease have not been explored. Extrapolating from our hospital-based wastewater data, it appears that peak fecal viral shedding may occur around symptom onset, given the congruence of wastewater RNA-signal and nosocomial cases and outbreaks identified in hospitals. Based on the rapid decline in wastewater signal thereafter, it is likely that fecal SARS-CoV-2-RNA drops significantly after initial presentation. Indeed, early in the pandemic when fewer patients with COVID-19 were hospitalized, wastewater samples routinely tested negative. This critical observation suggests that wastewater-based monitoring of SARS-CoV-2 may be most sensitive for identifying incident cases. Accordingly, wastewater-based monitoring of individual high-risk facilities (i.e., hospitals, nursing homes and industrial meat plants) – providing more granular data - through to WW-TP may have great merit.

A key limitation to the identification of SARS-CoV-2 in wastewater samples, relative to clinical samples (e.g., swabs) is the massive volume of water in which samples are diluted. This necessitates sample concentration. While procedures for the efficient recovery of non-enveloped viruses exist, researchers continue to search for satisfactory protocols for enveloped viruses such as SARS-CoV-2[27]. Many groups have explored ways to improve the sensitivity of wastewater SARS-CoV-2 detection. Diagnostic platforms with improved sensitivity and more impervious to impurities in the wastewater matrix (i.e., digital droplet RT-PCR) also show considerable promise[28, 29]. Sampling in the proximal sewershed may lead to day-to-day variance resulting in signal noise. This is particularly true for single-facility studies where potential extremes in individual virus shedding could confound results; limited data suggests significant variations in fecal viral load occur (from 10^3.4^ to 10^7.6^)[30]. This is a key challenge that remains to be solved if wastewater data is to be used in a meaningful way. Similarly, issues arise with attempts to normalize SARS-CoV-2 based on the contributing population. We chose to use the PMMoV as a fecal biomarker to control for variations in fecal loading – a particular risk when sampling takes place ‘upstream’ in the sewershed. While this marker has been validated in WW-TP samples where large and diverse populations contribute to sewage[31], hospitals are a much smaller collection of individuals and variations in PMMoV excretion owing to differences in diet could have a much larger impact[32].

By capturing longitudinal data from three tertiary-care hospitals (>2100 inpatient beds) we have demonstrated that passive wastewater monitoring is indeed possible at a range of hospital-facilities. Whereas Hospital-1 and Hospital-2 had a single municipal access point enabling surveillance of the entire facility – Hospital-3 required three locations to capture fully. As patients were frequently moved from one unit/building to another, either based on attending services geographic location, COVID-positive patient cohorting or need for ICU support, attributing SARS-CoV-2 signal in the wastewater of this facility was more complicated but nonetheless correlations were evident. Furthermore, while nursing staff is often assigned to individual units, many allied health workers and physicians work or consult throughout the entire facility. If passive wastewater monitoring is to be adapted for other aspects of nosocomial surveillance (i.e., antibiotic consumption, emergence of antimicrobial resistant organisms, etc.) – a keen insight into the collection network is required.

There are limitations of our work that merit discussion. Given the complexity involved in sample collection, we were limited to twice-weekly sampling. Knowing how quickly SARS-CoV-2 can spread and outbreaks can occur, a daily monitoring strategy would have much greater capacity to identify and mitigate secondary cases of COVID-19. Incident cases are likely to be clinically diagnosed in hospital far faster than in other high-risk facilities owing to greater resources and heightened suspicion – potentially leading to an even greater lead-time associated with a positive wastewater-signal of outbreaks than observed herein. Hospitals pose unique challenges in wastewater monitoring owing to these facilities’ high use of chemical disinfectants and detergents[33] that could interfere with molecular assays; this may explain why 3.6% of our samples spiked controls and PMMoV were not detected by RT-qPCR. To minimize the risk of false negatives, rigorous protocols that use internal controls (such as our BCoV spike) are necessary. The role of PCR-inhibitors in the wastewater matrix within the proximal sewershed is an area that deserves considerable study if this field is to expand.

Wastewater-based monitoring can only effectively monitor those individuals that contribute fecal matter to the sewershed. Importantly, hospitalized patients – those most vulnerable to COVID-19 adverse events – are often unable to self-toilet. Rather, these sick and often elderly individuals are dependent on continence aids, adult diapers, sanitary pads and nursing cleanup; this results in fecal matter from these individuals being disposed into biohazard solid waste. Accordingly, wastewater-based sampling could miss between 10-20% of patients in general hospital patients[34, 35]. This proportion is expected to be even higher in ICU where immobilization necessitated through ventilatory support further heightens toileting assistance requirements. Adapting wastewater surveillance technology to other high-risk settings like long-term care will encounter this same limitation. Finally, – tracking workers through passive wastewater monitoring poses inherent challenges. There is data that suggests that 52-56% of employees are uncomfortable defecating at work[36, 37].

## Conclusion

In a five-month observational study we were able to detect SARS-CoV-2 from the wastewater of Calgary’s three largest tertiary-care hospitals. The rate of SARS-CoV-2 wastewater test-positivity and RNA-abundance increased over time, concomitant with the increasing proportion of patients hospitalized with COVID-19. Despite persistent low levels of SARS-CoV-2-RNA in wastewater resulting from patients being treated for and recovering from COVID-19, we detected spikes attributable to hospital-acquired infections and outbreaks. This study reveals that wastewater-based monitoring of SARS-CoV-2 RNA holds promise for early detection, monitoring and containment of incident infections.

## Data Availability

Data will be made available after appropriate peer review when the manuscript is published.

## ACKNOWLEDGEMENTS

The authors gratefully acknowledge the staff of the City of Calgary and in particular members of Water Quality Services for their continued efforts for sample site planning, maintenance, and collection. The authors are grateful to staff from Alberta Health Services and AHS Infection Prevention and Control for assistance in data collection on patient disposition in the Calgary Zone. The authors also thanks Cameron Semper for providing the pMCSG53 vector and T7/T7 terminator primers, and for providing support with Gibson assembly cloning. We would like to acknowledge the tremendous efforts of Dr Rhonda Clark for program administration and management.

All authors report no potential conflicts. All authors have submitted the ICMJE Form for Disclosure of Potential Conflicts of Interest.

