## Supplementary material for "Wastewater Monitoring of SARS-CoV-2 from Acute Care Hospitals Identifies Nosocomial Transmission and Outbreaks": Online Supplment

### SUPPLEMENTARY METHODS

#### Acute care hospitals and hospital information systems

Calgary is Canada's fourth largest city, with a population of 1,286,000 residents in 2019 [1]. All hospital services in Calgary and the surrounding communities (total population = 1,670,000 residents) are provided by the publicly funded Alberta Health Services (AHS)– Calgary Zone, and care is delivered through four tertiary acute care adult hospitals and one acute care children's hospital. Together, these sites process 475,000 emergency room visits, a total 1,040,000 hospitalization-days and 145,000 hospital discharges –annually [2]. Sampling locations were chosen for their catchment size and ease of sanitary access, in consultation with the City of Calgary and Alberta Health Services (AHS). Three hospitals were studied encompassing 89% of regional tertiary-care adult beds: Hospital-1 (NE Calgary, 511 inpatient beds), Hospital-2 (SW Calgary, 611 inpatient beds) and Hospital-3 (NW Calgary, 1007 inpatient beds).

Prevalent cases were available only on the basis of the entirety of an individual hospital (i.e., unit-level data was not available) and prevalent data was available until December 3rd, 2020. Daily prevalent-hospitalized cases are recorded in an AHS Tableau Dashboard (Calgary Zone COVID-

19 Daily Activity) and represent patients with a RT-qPCR-confirmed COVID-19 diagnosis within 14 days and remain on contact/droplet precautions. After initially diagnosing nosocomial COVID-19 cases, we were not able to subsequently track the movement of patients or staff through hospital facilities (in particular at Hospital-3) given frequent patient movement from one unit to another, either based on attending services geographic location, or COVID-positive patient cohorting (i.e., in many instances cases on general medical/surgical wards were concentrated on a COVID-unit with special expertise) or transferred to the ICU owing to respiratory/hemodynamic worsening), attributing SARS-CoV-2 in the WW of Hospital-3 in particular, was much more complicated.

All cases attributable to a unit-specific outbreak were included in the total. This includes those cases that were diagnosed while still on that unit, those that had already been transferred to another unit, and those that had been discharged or transferred to another facility. Outbreaks involving HCW were defined if at least one of the HCWs was in the work-place during the communicable phase of illness or it is suspected there has been work site transmission as cause for one or more of the infections. When an outbreak was identified, all patients on the units were immediately placed on contact/droplet precautions, regularly screening and SARS-CoV-2 serial-testing of potentially exposed patients, enhanced cleaning, and closing the unit to new admissions to prevent subsequent spread and all potentially exposed HCW immediately removed from work for a minimum of 14 days of quarantine. Patients linked to outbreaks included those that were discharged home and subsequently diagnosed as occurring within 14 days of the outbreak, as well as those identified while still in hospital. Healthcare workers linked to outbreaks that developed RT-qPCR confirmed disease within 14 days of the event were identified using data extracted from AHS Workplace Health and Safety Regional Data Collection.

**Wastewater sampling**

ISCO GLS samplers were installed by highly-trained personnel from the City of Calgary at the designated manholes and programmed to collect 100 ml of wastewater every 15 mins, for a total of 96 samples over a period of 24 hours. These 24 hr composite samples were collected 2 times a week. Temperature readings of each samples were taken and recorded in the field, at the time of collection, as well in the laboratory during subsampling. Samples were transported to the City of Calgary's wastewater laboratory on ice. The 10 l carboy was well-mixed and poured off into five 500 ml sample bottles. Samples were stored at 4°C, in accordance with standard methods, before being shipped to the Advancing Canadian Wastewater Assets (ACWA) lab for analysis.

**Sample Processing and RNA Purification**

Each sample was spiked with 200 µl of Bovine coronavirus (BCoV) ($5 \times 10^5$ 50% tissue-culture-infective dose ($TCID_{50}$/ml)) positive control. BCoV aliquots were generated by resuspending a Bovilis® Coronavirus Vaccine dose (Merck, Catalogue #151921) in 2ml of PBS and stored at -80°C. Sample processing and RNA purification was conducted using the 4S (Sewage, Salt, Silica and SARS-CoV-2) method with a few modifications [3]. In brief, 40 ml aliquots of WW were transferred into 50 ml conical tubes where particle lysis and RNA preservation were conducted by the addition of 9.5 g of NaCl and 400 µl of TE buffer, respectively. The mixture was filtered through a 5 µm PVDF filter into 40 ml EtOH to remove large particles and debris. Subsequent RNA binding, washing, and elution was performed using a silica spin column (Zymo III-P silica spin column, Zymo Research) attached to a vacuum manifold. Washing volumes for buffers 4S-WB1 and 4S-WB2 were adjusted to 10 ml and 20ml respectively to minimize downstream inhibition. Nucleic acids were eluted in 100 µl of 50°C RNAse free water and stored immediately at -80 °C.

**RT-qPCR analysis**

All amplification reactions for SARS-CoV-2 detection were carried out with 5 µl of extracted RNA as initial template, and each reaction contained different concentration of primers and probes (Table 2) and 5 µl of TaqMan™ Fast Virus 1-Step Master Mix (Applied Biosystems). Reactions were performed at 50°C for 5 minutes, followed by polymerase activation at 95°C for 20 s, and 45 cycles of denaturation, annealing/extension at 95°C/3 s, then 55°C/3 s, respectively. Serial dilutions of the 2019-nCoV_N positive control (Integrated DNA Technologies, Catalogue # 10006625) and the 2019-nCoV_E positive control (Integrated DNA Technologies, Catalogue # 10006896) were run in triplicate on every 96-well PCR plate to produce standard curves used to quantify the copies of SARS-CoV-2 N1/N2 and E genes, respectively.

To estimate the number of genomic copies of PMMoV and the number of copies of the internal control (i.e., BCoV) in WW samples, a positive plasmid control was generated. Briefly, a sequence of double-stranded DNA fragment of 482 bp in length, containing the region 28938-29182 of the transmembrane (M) gene of the BCoV strain BCV-AKS-01 (GenBank: KU886219.1) and the region 1808-2040 of the replicase gene of the PMMoV isolate PMMoV-16.9 (GenBank: MN267901.1) was synthesized by Integrated DNA Technologies (IDT, Canada) and cloned into the pMCSG53 vector [4] via Gibson assembly. Inserts were confirmed by sequencing using the T7 (5'-TAATACGACTCACTATAG- 3') and T7 terminator (5'- GCTAGTTATTGCTCAGCGG -3') primers. The resulting plasmid was designated pBCoV-PMMoV. Amplification reactions for the internal and normalization controls detection were carried out with 4 µl of RNA as initial template, and each reaction contained different concentration of primers and probes (Table 2) and 5 µl of TaqMan™ Fast Virus 1-Step Master Mix (Applied Biosystems). Reactions for BCoV and PMMoV detection were performed at 50°C for 5 minutes, followed by polymerase activation at 95°C for 20 s, and 40 cycles of denaturation, annealing/extension at 95°C/3 s, then 60°C/3 s, respectively. Ten-fold dilutions of the pBCoV-PMMoV plasmid, representing 40 to $4x10^9$ copies of template were run in triplicate on every 96-well PCR plate to produce standard curves used to quantify the genomic copies of BCoV and PMMoV.

Genome copies per reaction were converted to copies per unit volume of wastewater (i.e., 40 ml) using the following dimensional analysis: Concentration (genomic copies/ml) = $C * V_{T\ eluted} * 1 / V_{per\ rxn} * 1/ V_{composite}$, where C = number of copies exported from the QuantStudio 5 Real-Time PCR System, $V_{T\ eluted}$ = total volume of nucleic acids eluted during silica column purification (µl), $V_{per\ rxn}$ = volume of nucleic acids added to each RT-qPCR reaction (µl) and $V_{composite}$ = volume of composite sample used to extraction (ml). To normalize the number of copies obtained for each of the three targets for SARS-CoV-2 detection (i.e., N1, N2 and E) by the detection of PMMoV in samples, we used the same approach described by D'Aoust *et al* [5].

To determine the limit of detection (LOD) of the N1 assay a similar approach described by La Rosa *et al* [6] was followed. Briefly, pure SARS-CoV-2 RNA was extracted from the NATtrolTM SARS-Related Coronavirus 2 stock (ZeptoMetrix, Catalogue # NATSARS(COV2)-ST) using TRIzol™ Reagent treatment and RNeasy Mini Kit (Quiagen, Catalogue # 74104) following the manufacturer's instructions. Then, the SARS-CoV-2 pure RNA (quantified ~$5 \times 10^3$ genome copies/ µl) was serial diluted to assess the sensitivity of the N1 assay. Similarly, to access the sensitivity in WW nucleic acid extracts, the same dilutions of the pure target were performed in nucleic acids extracted from a wastewater hospital sample that had tested negative for SARS-CoV-2. Each diluted samples was analyzed by quadruplicate and the LOD (reported as genome copies of SARS-COV-2 per ml wastewater) was determined as the last dilution where the relative repeatability standard deviation (RSDr) of the replicates was ≤ 33% [7].

**Sanger sequencing**

To confirm the signal detection obtained from the RT-qPCR analysis, nucleic acids samples with SARS-CoV-2 positive signal from two hospitals (i.e., Hospital-1 and Hospital-3) were subject to sequencing. Briefly, RNA samples were quantified using the DS-11 Quantification Spectrophotometer (DeNovix) and one microgram of total RNA was reverse transcribed using SuperScript IV Reverse Transcriptase (Invitrogen), according to the manufacturer's instructions. Then, each cDNA sample was used as template for amplifying a region of the nucleocapsid gene of SARS-CoV2 using the N_Sarbeco_F and N_Sarbeco_R primers [8]. PCR was performed with 5 µl of cDNA as template and with Platinum™ Taq DNA Polymerase (Invitrogen) with the following thermal cycling program: 94 °C for 2 min and then 35 cycles of 94°C for 30 s, 58°C for 30, and 72°C for 30. The PCR products were run on a 2% agarose gel and then purified, using Genomic DNA Clean & Concentrator kit (Zymo Research, Catalogue #: D4065), following the manufacturer's instructions and sent for sanger sequencing at the Sequencing facility of the University of Calgary, Canada.

### SUPPLEMENTARY RESULTS

#### Reproducibility of sample processing and RNA purification

The sampling timeframe included 18 weeks at Hospital-1 and Hospital-2, Hospital-3A and 11 weeks at each of Hospital-3B and Hospital-3C. To determine the reproducibility of sample processing among hospital samples, BCoV was used as an internal control for sample processing and RNA purification. Sample reproducibility was acceptable as similar BCoV concentrations in WW were observed among hospitals except for a few samples from Hospital-2 and Hospital-3A (see Supplementary Figure S1). The identified median number of spiked BCoV copies per ml of wastewater samples differed between hospitals (Hospital-1, $9.5 \times 10^5$ [IQR, $3 \times 10^5$– $2.1 \times 10^6$]; Hospital-2, $8.6 \times 10^5$ [IQR, $5.6 \times 10^5$– $2.1 \times 10^6$]; Hospital-3A, $2 \times 10^5$ [IQR, $6.7 \times 10^4$– $7.8 \times 10^5$]; Hospital-3B, $9.9 \times 10^5$ [IQR, $7.1 \times 10^5$– $1.5 \times 10^6$]; and Hospital3-C, $8 \times 10^5$ [IQR, $2.8 \times 10^5$– $1.2 \times 10^6$]; $P < 0.0001$). From the analysis, six samples were excluded owing to low BCoV signal (corresponding to samples collected on August 27th, September 3rd, 9th,10th and 17th from Hospital-3A and September 15th from Hospital-2). Furthermore, there was no detectable signal for the PMMoV human fecal biomarker in these excluded samples, suggesting issues with sample integrity (Figure S2). In general, the signal of BCoV recovery was lower from Hospital-3A location (Hospital-3A vs Hospital-3B, $P<0.0001$; Hospital-3A vs Hospital-3C, $P=0.0101$; Hospital-3A vs Hospital-1, $P= 0.0002$; Hospital-3A vs Hospital-2, $P <0.0001$).

Chemical analysis of water from Hospital-3A indicated significant spikes in chloride concentration, as high as 1780 mg/l from a baseline average of 100-200 mg/l at all other processing locations. Additionally, sample turbidity was visually noted to be unusually low for primary municipal wastewater, and samples were observed to have a free-chlorine smell. Correspondingly, total chemical oxygen demand averaged 173 mg/l in Hospital-3A samples compared to average anticipated values greater than 500 mg/l in typical primary municipal wastewater, indicating substantial oxidation of the wastewater prior to sample collection. Inspection of building schematics showed that Hospital-3A contained wastewater from a medical reprocessing facility within the hospital, likely causing the observed spike in chloride concentration as well as apparent free-chlorine. The presence of a strong oxidant in the form of chlorine bleach in samples from Hospital-3A presents a high probability of degradation of SARS-CoV-2 in the water, though the level of such degradation cannot be determined. Wastewater fecal strength signal (i.e., PMMoV) varied from 0.5 to 11015.2 genomic copies/ ml (median 182.2 copies/ml (IQR:42.5 - 841.6) among all hospitals (Figure S3). The median number of PMMoV copies per ml of sample differed between hospitals (Hospital-1, 133 [IQR, 48.6– 827]; Hospital-2, 143 [IQR, 43.7– 625]; Hospital-3A, 42.4 [IQR, 5.15– 241]; Hospital-3B, 1183 [IQR, 245– 2753]; and Hospital3-C, 429 [IQR, 72.9– 1383]; P <0.0001).

**SARS-CoV-2 Kinetics in hospital wastewater**

SARS-CoV-2 gene-targets were evaluated for their sensitivity. Relative to N1 (considered for this study the gold-standard), N2 and E had a sensitivity of 77.4% and 23.07 % respectively, – which did not differ across each site of sampling for N2 sensitivity (Hospital-3A: 80%, Hospital-3B: 81%, Hospital-3C: 66.7%, Hospital-1: 80% and Hospital-2: 77.8%, P>0.999), or E (Hospital_3A: 55.6%, Hospital-3B: 0%, Hospital-3C: 14.3%, Hospital-1: 27.3% and Hospital-2: 0%, P>0.999). The LOD for the N1-target was found to be 1.025 genome copies of SARS-COV-2 per ml wastewater.

Finally, we validated our RT-qPCR detection of SARS-CoV-2 from hospital wastewater samples by Sanger sequencing a 127 bp PCR product from the N-gene of two wastewater samples (i.e., Hospital-1: October 29$^{th}$ and Hospital-3A: September 29$^{th}$). The consensus alignment for the forward and reverse Sanger sequences for both samples analyzed confirmed a 100% identity match to the nucleocapsid phosphoprotein (N) gene of the SARS-CoV-2/human/CHN/Patient 12 isolate (GenBank: MW362756.1).

**SARS-CoV-2 in the city of Calgary during the study period**

During the period of observation, cases identified in the City of Calgary remained relatively stable from August at ~40-60 new cases/day (3.1 – 4.6 incident cases/100,000 residents per day) until mid-October when they began to increase, peaking in mid-December at ~600-700 cases/day (46.7-54.4 incident cases/100,000 residents per day). The absolute number of people in area hospitals on contact/droplet precautions generally mirrored community incident data (Figure S3).

### SUPPLEMENTARY TABLES

**Table S1.** Primers and probes for SARS-CoV-2 detection, spike detection and normalization control used in this study.

| Target | Assay Name | Oligonucleotide Sequence (5' – 3') | Final concentration | Reference |
|---|---|---|---|---|
| N gene | 2019-nCoV_N1-F | GACCCCAAAATCAGCGAAAT | 500 nM | [9] |
| | 2019-nCoV_N1-P | ACCCCGCATTACGTTTGGTGGACC | 125 nM | |
| | 2019-nCoV_N1-R | TCTGGTTACTGCCAGTTGAATCTG | 500 nM | |
| | 2019-nCoV_N2-F | TTACAAACATTGGCCGCAAA | 500 nM | |
| | 2019-nCoV_N2-P | ACAATTTGCCCCCAGCGCTTCAG | 125 nM | |
| | 2019-nCoV_N2-R | GCGCGACATTCCGAAGAA | 500 nM | |
| E gene | E_Sarbeco_F1 Forward Primer | ACAGGTACGTTAATAGTTAATAGCGT | 400 nM | [8] |
| | E_Sarbeco_P1 Probe | ACACTAGCCATCCTTACTGCGCTTCG | 200 nM | |
| | E_Sarbeco_R2 Reverse Primer | ATATTGCAGCAGTACGCACACA | 400 nM | |
| BCoV M gene | BCoV-F | CTGGAAGTTGGTGGAGTT | 200 nM | [10] |
| | BCoV-Pb | CCTTCATATCTATACACATCAAGTTGTT | 125 nM | |
| | BCoV-R | ATTATCGGCCTAACATACATC | 200 nM | |
| PMMoV replicase gene | PMMV-FP1-rev | GAGTGGTTTGACCTTAACGTTTGA | 400 nM | [11] |
| | PMMV-Probe1 | CCTACCGAAGCAAATG | 200 nM | [12] |
| | PMMV-RP1 | TTGTCGGTTGCAATGCAAGT | 400 nM | |

**Table S2.** RT-qPCR assays performance.

| Target | Number of plates | Y-intercept | Slope | Efficiency Range* | $R^2$ Range* |
|---|---|---|---|---|---|
| N1 | 16 | 36.1 – 38.6 | -3.6 - -2.4 | 90.7 – 161.7 | 0.7 – 1 |
| N2 | 16 | 38.7 – 46.2 | -4.1 - -3 | 75.4 – 117.2 | 0.9 – 1 |
| E | 9 | 36.2 – 41.3 | -4.1 - -3.1 | 75.2 – 111.1 | 0.8 – 1 |
| BCoV | 16 | 37.3 – 43.4 | -3.8 - -3.2 | 83.7 – 107.5 | 0.9 – 1 |
| PMMoV | 15 | 36.8 – 44.6 | -3.7 - -2.9 | 87 – 122.1 | 0.9 – 1 |

*Ranges are reported for all the plates ran for each target assay.

### SUPPLEMENTARY FIGURES

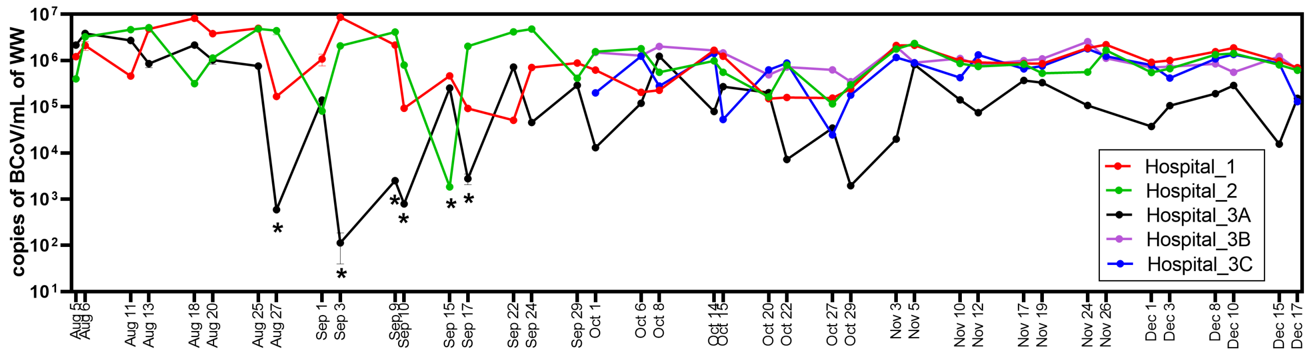


**Figure S1. Quantification of internal control in wastewater hospital samples.** Quantification of genomic copies of BCoV per ml of wastewater processed for 5 hospital locations over time determined by the transmembrane-protein gene of BCoV assay. The figure shows the average of three technical replicates in the $Log_{10}$ scale and error bars represent the standard deviation. Hospital-3B (purple) and Hospital-3C (blue) locations did not begin monitoring until October 1st. Asterisk (*) denotes the 6 samples that were excluded from the analysis as there was low BCoV signal and there was no detectable signal for the PMMoV human fecal biomarker.

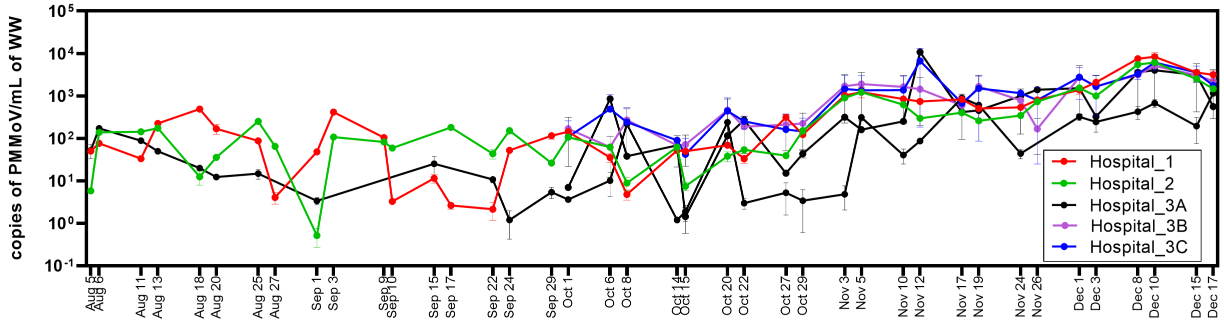


**Figure S2. Quantification of PMMoV normalization control in hospital wastewater samples.** Quantification of genomic copies of PMMoV per ml of wastewater processed for 5 hospital locations over time determined by the transmembrane-protein gene of BCoV assay. The figure shows the average of three technical replicates in the $Log_{10}$ scale and error bars represent the standard deviation. Hospital-3B (purple) and Hospital-3C (blue) began monitoring October 1st. Six samples corresponding to the following dates: August 27th, September 3rd, 9th,10th and 17th from location Hospital-3A and September 15th from location Hospital-2 were excluded from location Hospital-3A as there was not a detectible signal for PMMoV.

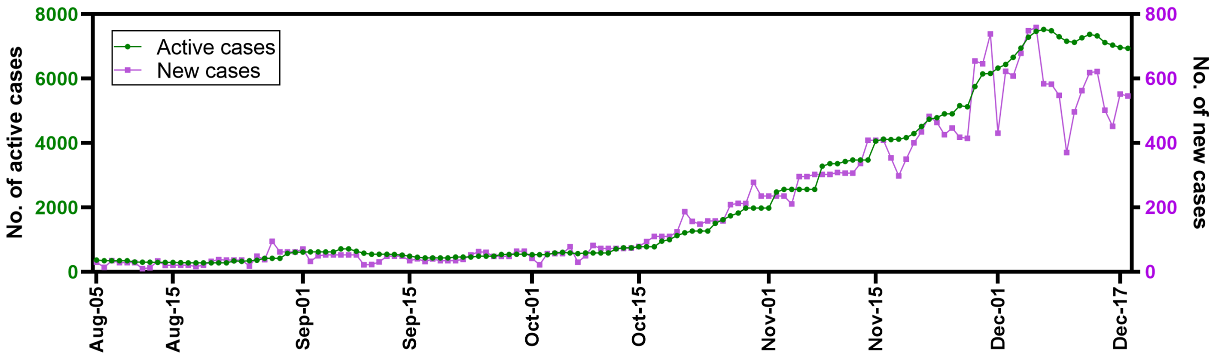


**Figure S3. Trends of the number of active and new cases of COVID-19 in the City of Calgary.** The figure shows the total number of active cases (green) and the number of daily confirmed cases (purple) for the City of Calgary during the time of this study (i.e., August 15th to December 17th, 2020). Epidemiological data was extracted from the Centre for Health Informatics (CHI) COVID-19 Tracker, Cumming School of Medicine, University of Calgary, Calgary.
